# Effect of a home-based health, nutrition, and responsive stimulation intervention and conditional cash transfers on child development and growth: a cluster-randomized controlled trial in Tanzania

**DOI:** 10.1101/2021.01.20.21250176

**Authors:** Christopher R. Sudfeld, Lilia Bliznashka, Geofrey Ashery, Aisha K. Yousafzai, Honorati Masanja

## Abstract

**Introduction:** Evidence on the effect of community health worker (CHW) interventions and conditional cash transfers (CCTs) on child growth and development in sub-Saharan Africa remains sparse.

**Methods:** We conducted a single-blind, cluster-randomized controlled trial of an integrated home-visiting health, nutrition, and responsive stimulation intervention alone and in combination with CCTs to promote antenatal and child clinic attendance from 2017 to 2019 in rural Morogoro region, Tanzania. Pregnant women and caregivers with a child <1□year of age were enrolled. Twelve villages were randomized to either a (i) CHW (n=200 participants), (ii) CHW+CCT (n=200), or (iii) control arm (n=193). An intention-to-treat analysis was conducted for the primary trial outcomes of child cognitive, language and motor development assessed with the Bayley Scales of Infant and Toddler Development and child length/height-for-age z-scores (HAZ) at 18-months of follow-up.

**Results:** The CHW and CHW+CCT interventions had beneficial effects on child cognitive development as compared to control (standardized mean difference (SMD): 0.15; 95% confidence interval (CI): 0.05, 0.24) and SMD: 0.18; 95% CI: 0.07, 0.28, respectively). The CHW+CCT intervention also had positive effects on language (SMD: 0.08; 95% CI: 0.01, 0.15) and motor development (SMD: 0.16; 95% CI: 0.03, 0.28). Both CHW and CHW+CCT interventions had no effect on HAZ in the primary analysis; however, there were statistically significant positive effects in multivariable analyses. The CHW+CCT group (mean difference: 3.0 visits; 95% CI: 2.1-4.0) and the CHW group (mean difference: 1.5 visits; 95% CI: 0.6-2.5) attended greater number of child health and growth monitoring clinic visits as compared to the control group.

**Conclusion:** Integrated CHW home-visiting interventions can improve child cognitive development and may have positive effects on linear growth. Combining CHWs with CCTs may provide additional benefits on clinic visit attendance and selected child development outcomes.

**Trial registration number:** ISRCTN10323949

**Key Questions Box:** *What is already known?:*

- Community health worker interventions that integrate health, nutrition and responsive stimulation components can improve child development but evidence from sub-Saharan Africa is limited.
- Conditional cash transfers can increase healthcare utilization but effects on child development and growth remain unclear.

*What are the new findings?:*

- An integrated home-visiting community health worker intervention benefited child cognitive development and may have improved child linear growth in rural Tanzania.
- Combining conditional cash transfers with the community health worker intervention increased child clinic visit attendance as intended and improved child cognitive, motor, and language development and may have improved child linear growth as compared to control.

*What do the new findings imply?:*

- Community health workers can improve child development and possibly child growth outcomes; however, additional research is needed to determine the intensity and frequency of visits to optimize impact as well as the direct and indirect mechanisms through which community health worker interventions work.
- Conditional cash transfers may provide additional benefits on clinic attendance and selected development domains as compared to community health workers alone; however, additional research is needed to directly compare integrated supply-side and demand-side strategies to promote child growth and development.

## INTRODUCTION

Community-based interventions that integrate health, nutrition and responsive stimulation components are a promising strategy to improve child health, growth and development.^1-3^ Home visit-based community health worker (CHWs) interventions primarily focused on health can increase the rate of facility births, uptake of child immunizations, and reduce newborn morbidity and mortality.^4 5^ Trials integrating responsive stimulation components in CHW interventions in low- and middle-income countries (LMICs) have also generally found positive effects on child development outcomes.^6-11^ Nevertheless, evidence on the effect of integrated child health, nutrition and responsive stimulation interventions is limited, particularly in sub-Saharan Africa. In addition, data on the effects of CHW interventions on child growth outcomes is sparse.^5 12^

While CHWs are a supply-side intervention providing additional demand-side conditional cash transfer (CCT) interventions to promote healthcare utilization have shown positive effects on maternal and child clinic visit attendance and child vaccination.^13^ However, evidence on the effect of CCTs on child nutrition, growth and development in LMICs is mixed, with the majority coming from Latin America.^13-15^ A recent meta-analysis showed that cash transfers have a small positive effect on height-for-age z-scores (HAZ) (difference HAZ: 0.03; 95% CI: 0.00, 0.05) and reduction of stunting (−2.1%; 95% CI: −0.69%, 3.5%) among children under 5 years of age; however there was no overall effect on weight-for-age z-scores (WAZ) and wasting.^16^ Further, a subgroup analysis found that the positive effects of cash transfers on HAZ was limited to studies in Asia and not studies in Latina America and sub-Saharan Africa, while positive effects on WAZ were found in studies in sub-Saharan Africa but not Asia and Latin America.^16^ In addition it was suggested that CCTs linked to a range of health, work, and education conditions had stronger effects on stunting as compared to unconditional cash transfers (UCTs).^16^ Nevertheless, the targeted age groups, size of the cash transfer, conditionalities, and delivery platforms varied widely between studies in the meta-analysis which makes the CCT and UCT evidence difficult to synthesize.

Cash transfers, with conditionality linked to parenting or educational programs, have generally shown greater impacts than unconditional cash transfers, including potential benefits on child development.^13^ Additionally, integration of parenting interventions (including responsive stimulation components) in CCT platforms has demonstrated positive effects on child development in Colombia ^17^ and Mexico^18^. However, in these studies, the CCT interventions were designed as a component of social protection program for poor families and were not directly designed to improve child growth and development. To the best of our knowledge, no study has directly targeted small CCTs to the general population of mothers and children in a community with conditionalities to increase access and utilization of antenatal care and routine child health growth monitoring to potentially promote child growth and development.

We present primary results from a trial that examined the effect of an integrated CHW-delivered health, nutrition, and responsive stimulation intervention alone and in combination with CCTs in rural Tanzania. We hypothesized that the CHW intervention would promote child growth and development through health and nutrition counselling, identification and referral for maternal and child illnesses, provision of early childhood development knowledge and promotion of caregiver responsiveness and developmentally appropriate play and communication activities. We hypothesized that integrating a CCT with the CHW intervention would increase access and utilization of antenatal and child health services, including child growth monitoring, treatment of health conditions, and other interventions not provided by the CHWs. The CHW and CCT interventions were designed in alignment within the program and resource-constraints in Tanzania to enhance the ability to translate the findings to scale. This proof-of-concept efficacy trial was intended to inform the need and design of larger effectiveness evaluations.

## METHODS

### Trial Design and Setting

We conducted a single-blind, cluster-randomized controlled trial of a home-based CHW-delivered health, nutrition, and responsive stimulation intervention alone and combined with CCTs to promote antenatal care and routine well child visits in rural Ifakara, Tanzania (trial registration: ISRCTN10323949). The full trial protocol is detailed elsewhere.^19^ This report presents the effect of the interventions on the primary outcomes of the study (endline child development and HAZ) and we also present the secondary anthropometric outcomes of weight-for-age z-scores (WAZ), and weight-for-height z-scores (WHZ). We plan to present the results for all other secondary trial outcomes in forthcoming reports.

The trial was conducted in 12 selected villages in the Ifakara Health Institute HDSS in Kilombero and Ulanga districts in Morogoro region.^20^ The Ifakara HDSS area is located approximately 450□km by road from Dar es Salaam and has a population of ∼□400,000 people. The HDSS area is predominately rural, and most residents are subsistence farmers. A recent study conducted among children 18-36 months of age in the study area found a 16.9% prevalence of low birthweight (<□2500□g) and a 36.2% stunting prevalence.^21^

### Trial Participants

The trial recruited pregnant women and mother/caregiver-infant pairs who lived in the study villages through a house-to-house survey. The inclusion criteria were: (1) permanent residence in a study village, (2) pregnant (self-reported) or had a child <□1□year of age at the enrollment visit, (3) and provide written consent. The exclusion criteria were: (1) enrollment in any other clinical trial or intervention study, or (2) child with severe physical or mental impairment. Potential participants were not aware of their village’s randomization arm at the time of seeking consent. In each study village, enrollment continued until all pregnant women and mothers/caregivers with a child <1 year of age were enrolled or until 50 participants were enrolled, whichever was reached first. If the mother had twins, one child was randomly selected for the trial and the same child was assessed at each time point. Written informed consent was obtained for all participants. Participants were referred to clinics at the time of enrollment and during outcome assessment visits if the research assistant identified an emergency maternal or child health issue or if the mother reported immediate risk of self-harm in the depression assessment.

### Randomization and Interventions

Village clusters were randomly allocated in a 1:1:1 ratio to one of the three trials arms: (i) CHW, (ii) CHW+CCT, or (iii) Control. The full details of the CHW, CHW+CCT and Control arms are described in the trial protocol and Supplementary Table 1.^19^ Briefly, the CHW intervention was the same in the CHW and CHW + CCT arms and therefore the CCT was the only difference between the two intervention arms. The control arm had access to the existing clinic-based maternal and child services (no CHW or CCT). Randomization was stratified by semi-urban (six villages) and rural (six villages) villages to increase the likelihood of baseline balance between arms. Randomization was done by a non-study statistician using a computer-generated randomization list with sequence blocks of three.

The same CHW intervention was delivered in the CHW and CHW+CCT arms. CHWs delivered an integrated health, nutrition, and responsive stimulation intervention in the home every 4-6 weeks for the trial duration of 18 months. Supplemental Table 1 presents a full description of the CHW intervention.^22^ Briefly, the trial CHWs received the year-long basic training on the national curriculum by the government before being hired by the project. The national CHW program was not implemented in the study area during the trial due to resource constraints, and the CHW salaries were paid by the project; the monthly salary for the CHWs was ∼□600,000 Tanzanian shillings (∼$230). Each CHW was assigned and delivered the intervention to two villages (∼100 participants). There was no CHW turnover over the course of the trial and therefore participants received the intervention from the same CHW for the duration of the trial. The CHW intervention included activities and duties of the standardized Tanzanian CHW curriculum with the addition of a responsive stimulation component (Supplemental Table 1).^19^ The intervention main maternal and child components of the CHW included: 1) identification and referral for under-5 childhood illness per Integrated Management of Childhood Illness (IMCI); 2) antenatal and postnatal pregnancy, delivery and essential newborn care counseling and danger signs identification; 3) family planning; 4) and emergency and routine referrals to facilities. The CHWs will be assigned to the study villages at the start of the trial. The CHWs provided counselling and referrals but did not directly provide treatments, medicines, nutritional supplements, immunizations or provide child growth monitoring. The Tanzanian government CHW curriculum does not include responsive stimulation or other direct early child development (ECD) promoting activities. The study team adapted the UNICEF and WHO Care for Child Development package to the local context and provided a one-week training in September 2017 on the responsive stimulation intervention that included integrated classroom and practical sessions.^23^ The responsive care component of the CHW intervention included essential early childhood development knowledge, promotion of caregivers’ sensitivity and responsiveness, and promotion of developmentally appropriate play and communication activities, toy making, and problem-solving. In addition, caregivers tried responsive stimulation activities with their young child and received feedback and coaching from the CHW. One field coordinator supervised CHWs through bi-weekly one-on-one meetings with each CHW, a monthly meeting with all CHWs, and monthly home visit spot-checks where the field coordinator accompanied CHWs to home visits (∼5% of visits).

A CCT intervention was also provided to participants in the CHW+CCT group every 4-6 weeks at the time of the CHW visit. The conditions for the CCT were attendance of routine antenatal care or routine well-child health and growth monitoring clinic visits. The CCT was intended to increase access and utilization of antenatal and child health services, including child growth monitoring, treatment of health conditions, and other interventions not provided by the CHWs. CHWs assessed antenatal care and child health cards at each home visit and provided mothers with cash payments of 10,000 Tanzanian Shillings ($4.30) per antenatal care visit or 5,000 Tanzanian Shillings ($2.15) per routine child health and growth monitoring visit that was completed since the last study visit. The average daily per person income for smallholder farmers in Tanzania is 1.90 USD.^24^ CHWs communicated that the CCT payments could be used in any way without penalty but suggested that mothers use the money for resources to support the health and development of the child.

### Assessments and Outcomes

Independent fieldworkers who were blinded to the randomized arm conducted home interviews with mothers at enrollment (baseline) and at 18 months after enrollment (endline). The fieldworkers were randomly assigned to villages each survey round and were not to ask participants about the intervention they received. The baseline visit occurred before implementation of the intervention, so it was not possible for the field workers to know the randomized group. Nevertheless, it cannot be ruled out that at the endline visit fieldworkers came to know the intervention status of a household from conversations with the mother. Standardized questionnaires were administered to collect demographic and socioeconomic data. In addition, the Hopkins Symptom Checklist (HSCL-25) was administered to assess symptoms of depression and anxiety^25^; symptoms consistent with depression were defined using the Tanzanian validated HSCL-25 cut-off.^26^ Functional social support was assessed using the Duke University– University of North Carolina Functional Social Support Questionnaire.^27^ The Caregiver Knowledge of Child Development Inventory (CKCDI) was administered at baseline.^28^ Child anthropometric measures were taken in triplicate in the home at the baseline and endline visit. Child weight was measured to the nearest 100 g using a digital scale (Seca, Hamburg, Germany). Child length (children < 24 months age) was measured to the nearest 0.1 cm using a length board (Seca, Hamburg, Germany) and child height was measured to the nearest 0.1 cm using a stadiometer (Seca, Hamburg, Germany). Anthropometric Z-scores were calculated using the 2006 WHO Child Growth Standards^29^. At endline, the fieldworkers administered process evaluations to each group. At endline, fieldworkers traveled outside the study area to conduct interviews and take anthropometric measurements for participants who moved temporarily or permanently outside the study area.

Female research nurses who were blinded to randomization arm administered a Tanzania adapted and Swahili translated version of the Bayley Scales of Infant and Toddler Development, Third Edition (BSID-III).^30 31^ The BSID-III was administered in quiet rooms at two health facilities serving the study area. The BSID-III nurses completed a three-week BSID-III training led by experts from Boston, USA and completed BSID-III assessments for a prior research study.^30^ For participants who moved outside of the study area, BSID-III assessments were not conducted. The two assessors differed in mean BSID-III domain composite scores (Supplemental Table 2) and therefore all analyses adjusted for assessor. However, BSID-III showed high internal consistency for all domains (Cronbach’s alphas ≥0.91) in the full sample and separately for each assessor (Supplemental Table 3).

### Sample Size

Sample size calculations were based on randomization of 12 village clusters, 50 mother/caregiver-child pairs per cluster, a nominal Type I error rate of 0.05 and an intra-cluster correlation of 0.01. We assumed 7.5% fetal loss or child death, 5% loss to follow-up (unknown vital status) and 15% missing data on LAZ. For child development, we originally planned to randomly select 60% of participants to have BSID-III assessed of which we assumed 10% would not complete the assessment (resulting in approximately 50%). Based on these assumptions, we had 80% power to detect a standardized mean difference (SMD) of 0.40 in length/height-for-age z-score (HAZ) and 0.53 standard deviations of development z-scores. However, to increase statistical power, we decided to invite all participants for BSID-III assessment. In a post hoc power analysis based on the actual number BSID-III assessments and observed within village correlation, we had 80% power to detect effect sizes of 0.48, 0.86, and 0.46 SD for cognitive, language and motor scores, respectively. The magnitude of the detectable differences for linear growth and development were large; however, this was a proof-of-concept efficacy trial and we hypothesized that there was potential of the intervention package and the combination of CHWs and CCT to provide a relatively large impact on child growth and development.

### Statistical Analysis

The intention-to-treat (ITT) principle was used for all primary analyses; participants who moved to neighboring villages were analyzed in the village originally randomized. All analyses accounted for clustering by village and urban/rural residence due to the stratified randomization scheme. Generalized linear regression models were used to assess the effect of the CHW and CHW+CCT interventions on the primary outcomes at endline: BSID-III sub-scale z-scores and HAZ. BSID-III z-scores were calculated using the internal mean and standard deviation. For the primary minimally adjusted analysis, BSID-III z-scores were adjusted for child age, sex, and BSID-III assessor; HAZ similarly adjusted for child age and sex. We also present BSID-III composite scores as a secondary outcome; however, applying US norms for BSID-III scores to children in other settings can result in misclassification and cross-cultural bias.^32 33^ The secondary outcomes of child weight-for-age z-scores (WAZ), weight-for-length/height z-scores (WHZ), and the number of child health and growth monitoring clinic visits were also assessed using generalized linear models. We did not analyze antenatal care visit attendance by randomized group since only 30% of women were pregnant at baseline and 60% of enrolled pregnant women delivered within 60 days of enrollment. Log-poisson models were used to examine relative risks of stunting (HAZ < −2), wasting (WHZ< −2), underweight (WAZ < −2) and overweight (WHZ > 2). The Benjamini-Hochberg procedure, which is a preferred method for accounting for multiple testing of correlated outcomes, was used to control for the potential false discovery rate for the eight primary analysis tests.^34 35^ Benjamini-Hochberg procedure adjusted p-values <0.05 were considered statistically significant.

We conducted sensitivity analyses that (i) adjusted for baseline factors which showed some degree of imbalance between randomization arms based on a p-value <0.20, and (ii) used stabilized inverse probability weights to account for dependent censoring (i.e. loss to follow-up).^36^ We also present effect estimates collapsing the CHW and CHW+CCT intervention arms. In addition, we explored the potential for effect modification by predefined baseline variables.^19^ The statistical significance of interaction was assessed with the likelihood ratio test and we did not adjust for multiple testing. Statistical analyses were performed with Stata Version 16 (StataCorp, College Station TX).

## RESULTS

Trial recruitment began in September 2017 and endline follow-up activities were completed in May 2019. In the 12 randomized clusters, 593 pregnant women or mother-infant pairs were recruited for participation and were analyzed by intention-to-treat. There were 50 participants in each of the four CHW and four CHW+CCT villages resulting in 200 participants in each group. There were 50 participants in two of the control villages and one control village with 47 participants and one with 46 participants which led to 193 total participants in the control group. Baseline characteristics were relatively comparable across intervention arms, but there was an indication of differences in household wealth, household sanitation, maternal education, parity, social support and CKCDI scores (Table 1 and Supplemental Table 4). The trial flow diagram is presented in Figure 1. The mean child age at endline assessment was 18.9 months (SD: 4.6). At endline anthropometric and BSID-III data was available for 91.5% and 67.7% of randomized participants, respectively. Children with endline anthropometric and BSID-III data were generally comparable at baseline to those without (Supplemental Tables 5 and 6). There were no adverse events reported during the trial.

**Table 1.** Baseline characteristics of trial participant stratified by randomized arm

|  | **Community Health**<br>**Worker**<br>**Mean ± SD or N (%)** | **Community Health**<br>**Worker + Conditional**<br>**Cash Transfer**<br>**Mean ± SD or N (%)** | **Control**<br>**Mean ± SD or N (%)** |
| --- | --- | --- | --- |
| Households/Mothers (n) | 200 | 200 | 193 |
| *Household characteristics* |  |  |  |
| Household size (persons) | 4.0 ± 1.7 | 3.6 ± 2.0 | 3.4 ± 1.7 |
| House has a dirt floor | 85 (42.5%) | 125 (63.5%) | 86 (44.6%) |
| Improved sanitation | 175 (87.5%) | 103 (51.5%) | 123 (63.7%) |
| Poorest wealth quintile | 28 (14.0%) | 50 (25.4%) | 46 (23.8%) |
| At least 1 toy in the home | 12 (8.3%) | 20 (23.8%) | 4 (3.3%) |
| *Maternal characteristics at baseline* |  |  |  |
| Age, years | 26.9 ± 5.3 | 27.0 ± 6.3 | 26.4 ± 6.1 |
| Married or living with partner | 172 (86.0%) | 167 (83.5%) | 149 (77.2%) |
| Education |  |  |  |
| No formal education | 8 (4.0%) | 36 (18.0%) | 8 (4.2%) |
| Primary education | 179 (89.5%) | 139 (69.5%) | 155 (80.3%) |
| Secondary or higher education | 13 (6.5%) | 25 (12.5%) | 30 (15.5%) |
| Pregnant at time of enrollment | 55 (27.5%) | 75 (37.5%) | 67 (34.7%) |
| Multiparous | 187 (93.5%) | 179 (89.5%) | 151 (78.2%) |
| Social support score | 2.1 ± 0.4 | 2.9 ± 0.6 | 2.9 ± 0.9 |
| Number of stimulation activities in the past 3 days | 1.6 ± 1.1 | 2.2 ± 0.8 | 2.5 ± 0.7 |
| Caregiver Knowledge of Child Development Inventory score (CKCDI) | 17.2 ± 4.4 | 13.8 ± 5.0 | 15.7 ± 5.2 |
| *Infant characteristics at baseline* |  |  |  |
| Infants (n; 0-1 year at enrollment) | 145 | 125 | 125 |
| Male | 75 (51.7%) | 70 (56.0%) | 62 (49.6%) |
| Age, months | 5.3±3.6 | 5.0±3.5 | 4.6±3.1 |
| Length-for-age z-score (LAZ) | -1.2±2.0 | -1.0±1.4 | 0.2±2.1 |
| Weight-for-length z-score (WLZ) | 0.0±1.1 | -0.3±1.3 | -0.2±1.4 |
| Weight-for-age z-score (WAZ) | 0.8±2.2 | 0.5±1.7 | -0.4±2.3 |

SD = standard deviation

**Figure 1.**
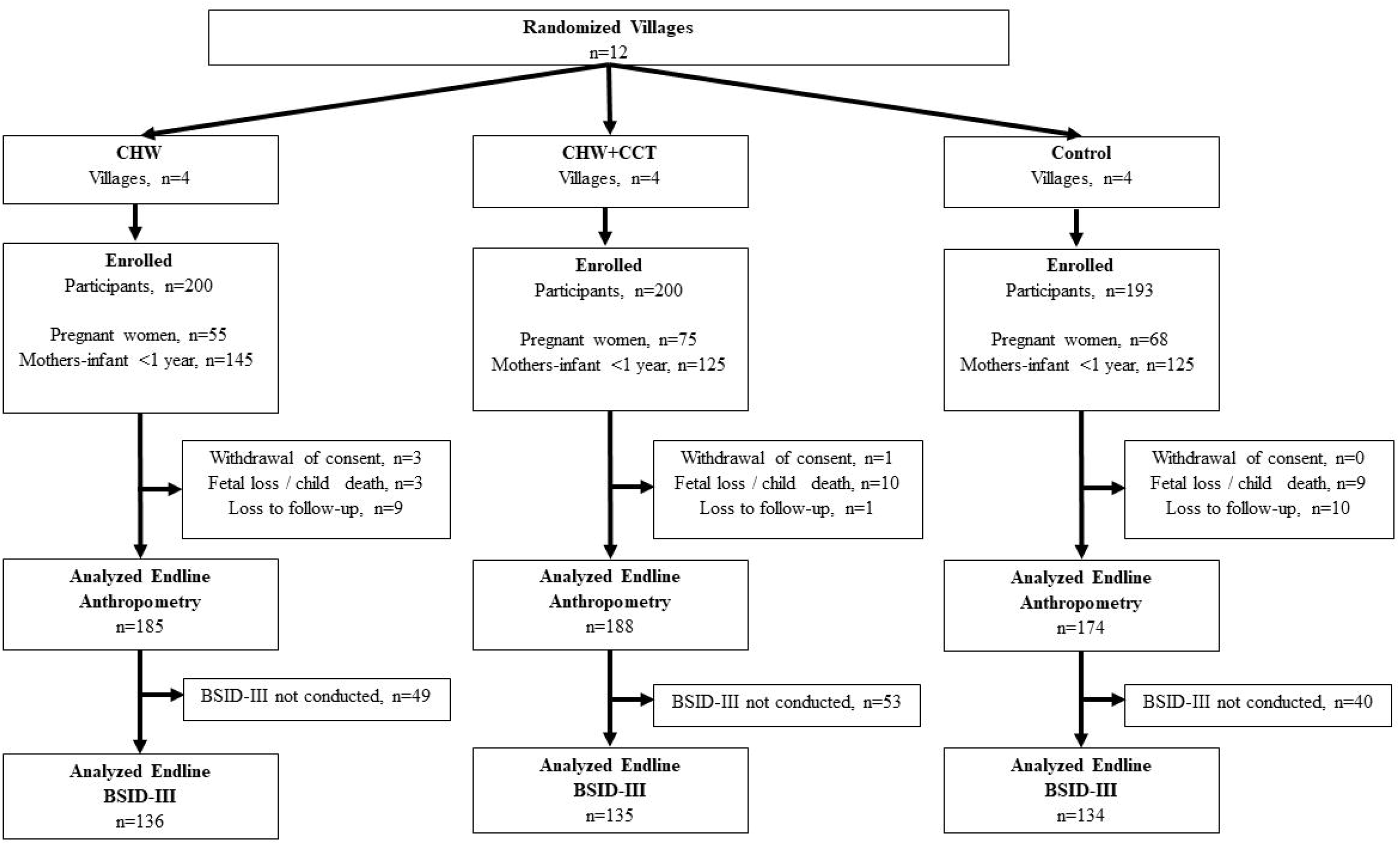
Trial flow diagram stratified by randomized arm.

The CHW and CHW+CCT interventions were delivered every 4-6 weeks as expected. The median number of home visits for the CHW arm was 11 (Q1: 9, Q3:13) and for the CHW+CCT arm was 12 (Q1: 10, Q3: 13). The percentage of the total expected CHW visits completed in the CHW group was 93% and 99% in the CHW + CCT group. The mean CHW visit duration in both intervention groups was approximately 35 minutes (mean 34.4 ± 3.3 CHW group and 35.2 ± 3.5 minutes CHW+CCT group). Process indicators and maternal opinions on the CHW and CCT interventions are presented in Table 2. Overall, these data suggest that coverage and fidelity of the CHW and CCT interventions was high. Further, we evaluated clinic visit attendance as an important intermediary as it was the condition for the cash transfer. Participants in the CHW+CCT attended 3.0 (95% CI: 2.1-4.0) more child health and growth monitoring clinic visits as compared to control, while participants in the CHW arm attended 1.5 additional clinic visits (95% CI 0.6-2.5) (Supplemental Table 7).

**Table 2.**
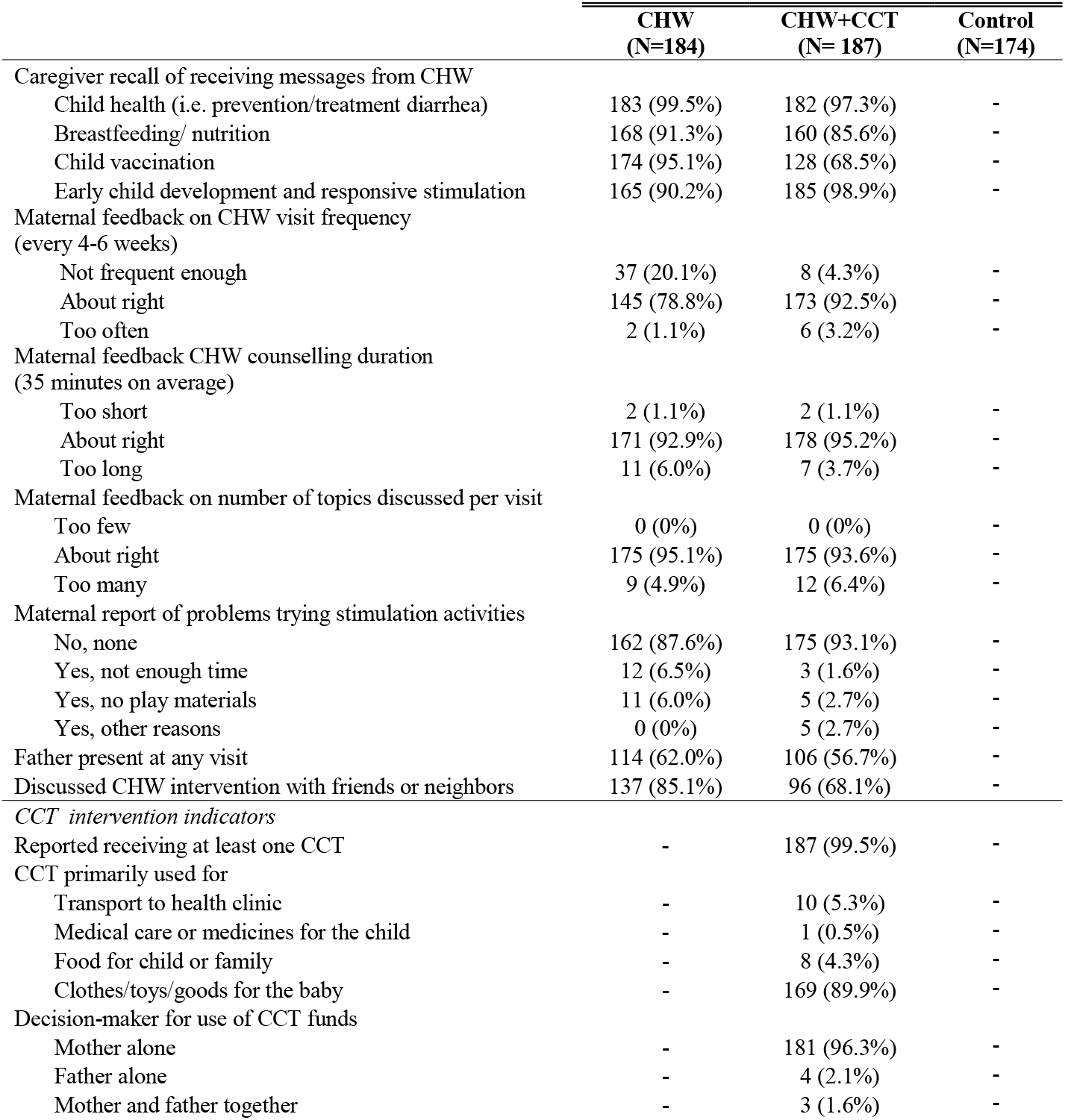
Process indicators and maternal feedback on community health worker (CHW) and conditional cash transfer (CCT) interventions

|  | CHW<br>(N=184) | CHW+CCT<br>(N= 187) | Control<br>(N=174) |
| --- | --- | --- | --- |
| Caregiver recall of receiving messages from CHW |  |  |  |
| Child health (i.e. prevention/treatment diarrhea) | 183 (99.5%) | 182 (97.3%) | - |
| Breastfeeding/ nutrition | 168 (91.3%) | 160 (85.6%) | - |
| Child vaccination | 174 (95.1%) | 128 (68.5%) | - |
| Early child development and responsive stimulation | 165 (90.2%) | 185 (98.9%) | - |
| Maternal feedback on CHW visit frequency<br>(every 4-6 weeks) |  |  |  |
| Not frequent enough | 37 (20.1%) | 8 (4.3%) | - |
| About right | 145 (78.8%) | 173 (92.5%) | - |
| Too often | 2 (1.1%) | 6 (3.2%) | - |
| Maternal feedback CHW counselling duration<br>(35 minutes on average) |  |  |  |
| Too short | 2 (1.1%) | 2 (1.1%) | - |
| About right | 171 (92.9%) | 178 (95.2%) | - |
| Too long | 11 (6.0%) | 7 (3.7%) | - |
| Maternal feedback on number of topics discussed per visit |  |  |  |
| Too few | 0 (0%) | 0 (0%) | - |
| About right | 175 (95.1%) | 175 (93.6%) | - |
| Too many | 9 (4.9%) | 12 (6.4%) | - |
| Maternal report of problems trying stimulation activities |  |  |  |
| No, none | 162 (87.6%) | 175 (93.1%) | - |
| Yes, not enough time | 12 (6.5%) | 3 (1.6%) | - |
| Yes, no play materials | 11 (6.0%) | 5 (2.7%) | - |
| Yes, other reasons | 0 (0%) | 5 (2.7%) | - |
| Father present at any visit | 114 (62.0%) | 106 (56.7%) | - |
| Discussed CHW intervention with friends or neighbors | 137 (85.1%) | 96 (68.1%) | - |
| *CCT intervention indicators* |  |  |  |
| Reported receiving at least one CCT | - | 187 (99.5%) | - |
| CCT primarily used for |  |  |  |
| Transport to health clinic | - | 10 (5.3%) | - |
| Medical care or medicines for the child | - | 1 (0.5%) | - |
| Food for child or family | - | 8 (4.3%) | - |
| Clothes/toys/goods for the baby | - | 169 (89.9%) | - |
| Decision-maker for use of CCT funds |  |  |  |
| Mother alone | - | 181 (96.3%) | - |
| Father alone | - | 4 (2.1%) | - |
| Mother and father together | - | 3 (1.6%) | - |

CHW and CHW+CCT intervention effects on BSID-III domain scores are presented in Table 3. In the primary analysis, both the CHW and CHW + CCT arms had beneficial effects on cognitive development scores and the CHW+CCT arm had positive effects on language and motor scores (Benjamini-Hochberg procedure p-values <0.05). The effect sizes were similar, and the findings were qualitatively the same in multivariable models that adjusted for potential baseline imbalance (Table 3). The results for analyses of BSID-III composite scores were similar to the primary analyses of BSID-III z-scores; the CHW and CHW + CCT arms had beneficial effects on cognitive composite scores and the CHW+CCT arm had positive effects on language and motor composite scores (Supplemental Table 8). In addition, the results were materially unchanged in a sensitivity analysis using inverse probability weighting accounting for dependent censoring (Supplemental Table 9). In secondary analyses, collapsing the CHW and CHW+CCT arms, there was a positive effect on cognitive scores but no effect on language or motor scores (Supplemental Table 10). We also explored potential modifiers of the effect of the interventions on BSID-III scores. The CHW+CCT intervention appeared to provide greater positive effects on language scores for infants whose mothers had lower baseline CKCDI scores and on motor scores for mothers with less than secondary education (p-values for interaction <0.05) (Supplemental Table 11).

**Table 3.** Effect of Integrated Community Health Worker Intervention (CHW) and CHW plus conditional cash transfer (CCT) on standardized mean difference in Bayley Scales of Infant Development Scores –III at 18 months of follow-up

|  |  |  |  | Primary minimally adjusted analysis<sup>a</sup> |  |  |  | Multivariable adjusted analysis<sup>b</sup> |  |
| --- | --- | --- | --- | --- | --- | --- | --- | --- | --- |
|  | CHW<br>Mean ± SD<br>(N=136) | CHW+CCT<br>Mean ± SD<br>(N=135) | Control<br>Mean ± SD<br>(N=134) | CHW vs.<br>Control<br>Mean Difference<br>(95% CI) | p-value<sup>c</sup> | CHW+CCT vs.<br>Control<br>Mean Difference<br>(95% CI) | p-value<sup>c</sup> | CHW vs. Control<br>Mean Difference<br>(95% CI) | CHW+CCT vs.<br>Control<br>Mean Difference<br>(95% CI) |
| Cognitive | 0.10 ± 1.12 | 0.01 ± 0.85 | -0.10 ± 1.01 | 0.15 (0.05, 0.24) | 0.009 | 0.18 (0.07, 0.28) | 0.008 | 0.14 (0.05, 0.22) | 0.14 (0.07, 0.21) |
| Language | 0.07 ± 1.06 | -0.13 ± 0.93 | 0.06 ± 1.00 | 0.04 (-0.06, 0.14) | 0.48 | 0.08 (0.01, 0.15) | 0.04 | 0.03 (-0.07, 0.13) | 0.07 (-0.02, 0.17) |
| Motor | 0.03 ± 1.08 | 0.04 ± 0.90 | -0.07 ± 1.02 | 0.04 (-0.08, 0.16) | 0.53 | 0.16 (0.03, 0.28) | 0.03 | 0.04 (-0.08, 0.16) | 0.18 (0.01, 0.35) |

<sup>a</sup>Minimally adjusted model included covariates for child age at assessment, sex, and BSID-III assessor and accounted for clustering
<sup>b</sup>Multivariate model included covariates for urban/rural residence, baseline household wealth quintile, household having access to an improved latrine, maternal education, parity, social support, CKCDI, child sex, child age at assessment, sex, and BSID-III assessor and accounted for clustering
<sup>c</sup>Benjamini-Hochberg procedure adjusted p-values

Intervention effects on child growth is presented in Table 4. In the primary analysis, there was no statistically significant effect on HAZ for either the CHW (mean difference HAZ: 0.83; 95% CI: −0.56, 2.22; Benjamini-Hochberg procedure p-value = 0.32) or CHW+CCT interventions (mean difference HAZ: 1.40; 95% CI: −0.04, 2.84; Benjamini-Hochberg procedure p-value = 0.09). However, in multivariable models there were significant beneficial effects on HAZ in both the CHW and CHW+CCT arms as compared to control. Further, the results were similar in a sensitivity analysis accounting for dependent censoring (Supplemental Table 12). Secondary analyses of the collapsed CHW and CHW+CCT arms also found a significant beneficial effect on HAZ in the multivariable model (Supplemental Table 13). In exploratory analyses of potential effect modifiers (Supplemental Table 14), there was consistent evidence that the magnitude of the positive effects of both the CHW and CHW+CCT interventions on HAZ was greater for women with lower social support (p-values for interaction <0.01). There was also indication that the CHW intervention provided greater benefits on HAZ for mothers with baseline depression and for mothers over 25 years of age as compared to younger mothers (p-values for interaction <0.05). There was no indication of a difference in the effect of either the CHW or CHW + CCT intervention HAZ if the mother was pregnant at enrollment or had a child <1 year of age (p-values >0.05). We also conducted a sensitivity analysis that examined the effect of the CHW and CHW+CCT interventions on child growth outcomes among the subgroup of participants that were <1 year at enrollment and additionally adjusted for their baseline HAZ in multivariable models due to potential for baseline imbalance (Supplemental Table 15); the positive effects of both the CHW (mean difference 0.74; 95% CI: 0.05, 1.42) and CHW + CCT groups (mean difference 1.35; 95% CI: 0.67, 2.03) on HAZ were significant in the multivariable analysis and similar in magnitude to the effect seen in the full trial population.

**Table 4.**
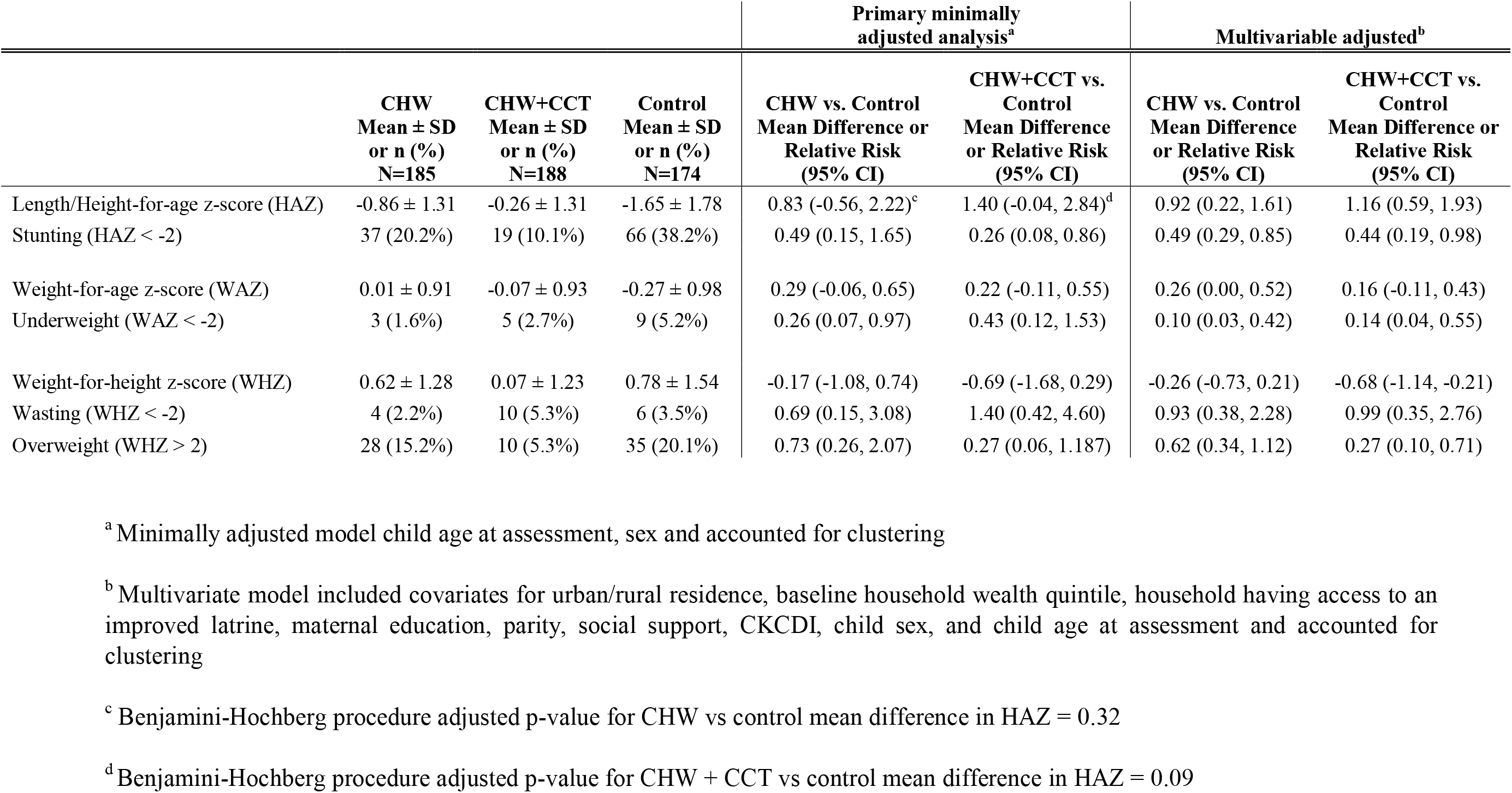
Effect of the integrated Community Health Worker (CHW) intervention and CHW plus conditional cash transfer (CCT) intervention on anthropometric outcomes at 18 months of follow-up

As for secondary anthropometric outcomes, the CHW arm reduced the risk of underweight and the CHW+CCT intervention reduced the risk of overweight in multivariable models (Table 4). The effect CHW on WAZ was greater for mothers ≥25 years as compared to <25 years, and for participants with poorer household wealth (<50th percentile) as compared to richer households (≥50th percentile) (Supplemental Table 14). The effect of CHW+CCT on WAZ appeared larger for women who were pregnant at baseline as compared to those that had a child <1 year at enrollment, participants with poorer household wealth (<50th percentile) as compared to richer households (≥50th percentile), and for mothers with lower social support as compared to higher social support (Supplemental Table 14). The effect of CHWs on WHZ was greater for mothers ≥25 years as compared to <25 years, and for mothers without baseline depression as compared to mothers with baseline depression (Supplemental Table 14).

## DISCUSSION

In this single-blind, cluster-randomized controlled trial conducted in rural Tanzania, we found that both the CHW and CHW+CCT interventions had beneficial effects on child cognitive development. The CHW+CCT arm also had positive effects on language and motor development. In addition, we found no statistically significant effect of either intervention on child HAZ in the primary analysis; however, both the CHW and CHW+CCT interventions had beneficial effects on HAZ in multivariable analyses.

We found positive effects of the CHW and CHW+CCT interventions on child cognitive development and this finding is in line with evidence on CHW home visit interventions that integrate health, nutrition and responsive stimulation interventions which have generally found moderate positive effects on child development outcomes^7^A meta-analysis of 21 such interventions determined positive pooled effect sizes of 0.42 SD and 0.47 SD for cognitive and language development, respectively.^6 7^ The Nurturing Care Framework notes that multiple interrelated components, including health care, adequate nutrition, responsive caregiving, opportunities for early learning, and safety and security create an enabling environment that promote child growth and development.^37^ CHW interventions may be able to address each of these components to varying degrees and therefore may provide greater impact on child growth and development than interventions which focus on individual components, such as health, nutrition, or stimulation interventions alone. Nevertheless, in our trial, we found relatively small beneficial effects of both the CHW and CHW+CCT interventions on cognitive development scores; however, the magnitude of the effect was only 0.1 SD; the CHW+CCT arm also had similar magnitude effects of 0.1 SD on language and motor scores. One potential explanation for the smaller effect size is that our trial provided a less intensive intervention by design as compared to prior trials and studies. In our study, CHW home visits were conducted every 4-6 weeks for an average of 35 minutes. Prior trials have found relatively large positive effects with 30-60 minute home visits; however, most conducted home visits every two weeks or more frequently.^7^ A trial of an integrated responsive stimulation and nutrition intervention implemented in rural Pakistan also conducted monthly home visits and group sessions and found positive effects on child cognitive, language and motor scores at 24 months of age but the group sessions were almost three times longer at 1 hour and 20 minutes.^38^ As a result, combining CHW interventions with group meetings or other supplemental activities that bolster behavior change may be important to enhance effects. It is also important to note that while the responsive stimulation component of the CHW intervention was designed to directly promote child development it is possible that the health, nutrition, and other support that CHWs provided may have contributed to the positive effects on child development in our trial per the Nurturing Care Framework.^37^ Prior trials, including the Pakistan trial^38^, that compared CHWs with responsive stimulation versus CHWs alone do not capture the potential benefits of CHWs through health, nutrition, and other factors..^6 7^ Nevertheless, the effect of CHW and CHW + CCT versus control in our trial was similar, if not smaller, than trials that compared CHW with responsive caregiving to CHWs without responsive caregiving. Additional research is needed to understand the direct and indirect mechanisms through which CHW intervention components may provide an enabling environment for positive effects on child development.

The CHW+CCT arm had similar magnitude of effect on cognitive and language scores relative to the CHW arm (although the effect on language was only statistically significant in the CHW+CCT arm). However, there was some suggestion, although not definitive, that the effect on motor development may be larger for CHW+CCT than CHW alone. In disadvantaged communities, toys and manipulatives that may enhance complex cognitive, language and motor skills are often scarce. Chang et al distributed a picture book and a puzzle (key examples of learning materials that support complex developmental skills) at well child visits to support a health-center based stimulation program in Jamaica that found positive effect on child cognitive development.^39^ In our study, the majority of mothers in the CHW+CCT reported that the CCT funds were used directly for goods for the infants, including toys, although social desirability may affect these findings. Several previous interventions encourage making homemade toys or sharing toys in communities over the course of the program; however, research on provision of toys and manipulatives targeting complex or higher-order skills development in disadvantaged and resource-scarce settings should be pursued alongside supporting responsive caregiver-child interactions and play.

In addition, the CHW and CHW+CCT interventions may have had positive effects on linear growth. It is important to note these positive effects were only statistically significant in multivariable models and that even though the point estimates indicate large effects the confidence intervals are wide and indicate that moderately small to very large beneficial effects are possible. There are multiple components of the CHW intervention evaluated in our trial that may have contributed to improvements child linear growth, including nutritional counselling, infection prevention and control, identification and referral for childhood illnesses, promotion of growth monitoring attendance, and responsive caregiving.^4 5 40^ A recent systematic review determined that CHW home visits increased early initiation of breastfeeding and exclusive breastfeeding.^4^ Provision of education on complementary feeding has also been shown to increase HAZ by 0.29 SD.^40^ Further, community health worker interventions may reduce incidence of childhood illnesses, including diarrhea.^5 41 42^ The CHW intervention evaluated in our trial was comprehensive by design and we cannot untangle which components or pathways may have led to improvement in linear growth. It is important to note that CHWs in our trial completed the one-year Tanzania government certified CHW training program, which is significantly longer training duration and more comprehensive in terms of interventions to be delivered in the home than other CHW programs, which may have resulted in beneficial effects. We did not find the effect of the CHW or CHW+CCT on growth or development differed for participants that were pregnant at baseline as compared to participants that were mothers and children <1 year of age at baseline. As a result, it is not clear if providing the interventions in pregnancy and postnatal period is greater than the postnatal period alone.

The CHW+CCT arm appeared to provide beneficial effects on linear growth that were similar in magnitude to the CHW arm in multivariable analyses. Evidence on the effect of cash transfers on child nutritional status is mixed, but there is relatively consistent evidence of positive effects on child diet and morbidity.^15^ The framework by Smith and Haddad suggests that CCTs may effect child nutritional outcomes through three main pathways of food security, health and access to care.^43^ We found that the CHW+CCT increased child clinic visit attendance as compared to control, which is consistent with evidence that cash transfers can be used to increase health service utilization in LMICs.^13 44^ Nevertheless, there remains a debate on the actual effectiveness of growth monitoring programs to provide beneficial effects on child growth.^45^ Further, it seems unlikely that the small cash transfers in our trial (4.30 USD per antenatal care visit and 2.15 USD per monthly child visit) would consistently improve food security. There is some evidence that cash transfers of 15% to 25% of total monthly household income in the context of a social protection program may provide greater effect on child nutritional outcomes; however, in our trial the relatively monthly cash transfers conditioned on clinic visit attendance were <10% of total monthly household income for subsistence farmers in Tanzania.^24 46^ As a result, additional research is needed to determine the amount and if and which conditionalities for cash transfers may provide beneficial effects on child growth and development.

This study has several important limitations. First, due to the small number of clusters and participant sample size; there was an inherent risk of baseline imbalances. However, multivariable analyses including adjustment for potentially imbalanced factors resulted in similar point estimates to the minimally adjusted models which suggests low risk of bias due to baseline imbalance. Nevertheless, the standard errors and confidence intervals were smaller in multivariable models as compared to minimally adjusted models, in particular the standard errors and confidence intervals were roughly one-half the size in the multivariable LAZ/HAZ analysis. It is well-documented in randomized trials that adjusting for prognostic baseline covariates that are associated with outcomes of interest can substantially increase statistical power by explaining variation between participants and not bias results^47 48^, some suggest randomized trials should routinely adjust for prognostic covariates to increase statistical power in the primary analysis.^49^ Second, our analyses of potential effect modifiers, although prespecified, were at risk of type I errors due to multiple testing and these results should be used for hypothesis generation. Third, BSID-III data was available for 67% of participants and therefore there is a risk of selection bias; however, there appeared to be no difference between children who were assessed versus not assessed for development outcomes. Fourth, the effects of the interventions were evaluated after 18 months of delivery and therefore it is not clear if the impact is sustained later in childhood.

Further, we did not have a CCT only arm and were therefore not able to directly assess interaction between CHW and CCT interventions. In addition, we designed our trial to compare the CHW intervention which included responsive stimulation compared to control (no CHW) to capture the full potential effects of the CHW package on child growth and development, as a result, we are not able to isolate whether the responsive stimulation component provided additional benefit beyond that of the CHW program alone. Further, we collected data on process indicators for the CHW and CCT interventions, but we did not have data on the counselling and interventions that participants received at each antenatal or child clinic visit and are therefore not able to assess quality of care in the clinics. However, we expect that the quality of care to be similar across groups due to randomization. Given endline data collection was collected 18 months and not every month, it is likely that maternal report of the services provided at each of the multiple clinic visits during the past 18 months would be highly prone to recall bias.

Nevertheless, our findings on the effect of the CHW and CHW+CCT interventions are applicable to the current context of clinic care in rural Tanzania and similar settings. Last, in our trial intervention delivery was carefully monitored and therefore determined efficacy of the interventions and consequently the effectiveness of the interventions in large-scale programs needs to be evaluated.

## Conclusions

Implementation research is needed to determine how to best integrate responsive stimulation components into CHW programs considering programmatic constraints that can vary by country, program, and context. Future studies should evaluate the cost-effectiveness of integrating responsive stimulation into CHW programs at scale. Our study also suggested that there may be beneficial effects of integrating CHWs with CCTs on some outcomes but additional research on integrated supply- and demand-side strategies to promote child growth and development is needed.

## Data Availability

Deidentified individual participant data (including data dictionaries) may be made available, in addition to study protocols, the statistical analysis plan, and the informed consent form. The data will be made available upon publication to researchers who provide a methodologically sound proposal for use in achieving the goals of the approved proposal and obtain ethical approval. Proposals should be submitted to.

## Acknowledgements

We would like to acknowledge the study staff and the all the mothers, in and for making the study possible.

## Contributors

CRS and HM conceptualized the study. CRS, LB, GA, AKY, and HM developed the intervention and trial protocol, GA coordinated, and supervised data collection, MB and CRS conducted the statistical analysis. CRS wrote the first draft of the paper. All authors reviewed and contributed to the draft paper. All authors approved the final submission.

## Patient and public involvement

Patients and/or the public were not involved in the design, or conduct, or reporting, or dissemination plans of this research.

## Patient consent for publication

Not required.

## Ethics approval

The trial protocol was approved by the Ifakara Health Institute IRB (reference number 007–2017), the Tanzanian National Health Research Ethics Sub-Committee (NatHREC; reference no. NIMR/HQ/R.8a/Vol.IX/2538), and the Harvard T.H. Chan School of Public Health Institutional Review Board (reference no. IRB17–1001).

## Provenance and peer review

Not commissioned; externally peer reviewed.

