## Supplemental Tables for "Effect of a home-based health, nutrition, and responsive stimulation intervention and conditional cash transfers on child development and growth: a cluster-randomized controlled trial in Tanzania"

**Supplemental Table 1.** Template for Intervention Description and Replication (TIDieR) table for the CHW home-delivered integrated health, nutrition, and responsive stimulation intervention that was delivered with the same strategy in the CHW and CHW+CCT groups.

| **Name** | CHWs delivered integrated health, nutrition and responsive stimulation intervention |
| --- | --- |
| **Why** | Intervention packages that include multiple health, nutrition, and responsive stimulation components may be impactful in promoting children’s growth and development. Community-based and integrated strategies may also provide greater intervention coverage, impact, and address the limited human and financial resources in low- and middle-income countries (LMIC) |
| **What** | The health and nutrition intervention components were directly aligned with the Tanzanian certified CHW program and included 1) identification and referral for under-5 childhood illness per Integrated Management of Childhood Illness (IMCI); 2) antenatal and postnatal pregnancy, delivery and essential newborn care counseling and danger signs identification; 3) family planning; 4) and emergency and routine referrals to facilities. CHWs provided nutrition counseling on maternal nutrition during pregnancy and lactation, and counseling on infant and young child feeding practices including exclusive breastfeeding to 6 months, continued breastfeeding to 24 months and safe, appropriate and adequate complementary feeding; CHWs also reviewed maternal and child health cards at each visit to inform counselling. The responsive stimulation component was a Tanzanian and Swahili adapted version of the UNICEF and WHO Care for Development package. The intervention included communication of essential early childhood development knowledge, promotion of caregivers’ sensitivity and responsiveness, developmentally appropriate play and communication activities and problem solving. In addition, CHWs provided advice on toy-making using items commonly found in the home and environment; no toys or play materials were directly provided to participants. Caregivers were encouraged to use everyday items in the home (e.g., cups for stacking), explore and talk about the home and surrounding natural environment and make playthings.  A field coordinator provided supervision of the CHWs throughout the intervention. The field coordinator had a Bachelor of Arts degree and research experience. Supervision included one-on-one biweekly meetings with each CHW, a monthly meeting with all CHWs, as well as monthly home visit spot-checks where the field coordinator accompanied CHWs during home visits. |
| **Who provided** | Female CHWs who resided in the study area. All CHWs completed secondary school education and were caregivers for children. |
| **Training** | The government CHW curriculum included two semesters that each covered seven topics. The first semesters covered: (1) fundamentals of communication and customer service, (2) infection prevention and control, (3) management of health care facility, (4) computer application, (5) citizenship and gender, (6) management information systems, and (7) basic life support skills. The second semester covered: (1) fundamentals of social work, (2) disease prevention and control, (3) community-based reproductive, maternal and child health services, (4) community-based health promotion, (5) home-based care, (6) basics of entrepreneurship and life skills, and (7) health facility and community disease management. For the responsive stimulation component, the CHWs received a one-week classroom-based intervention- specific training prior to the start of the intervention. This training covered both theoretical and practical aspects of early child development, age-appropriate play and communication activities with coaching techniques to prompt and guide caregiver’s responsiveness during the interaction, counseling of caregivers, problem solving, and making of toys and other play materials. A three-day refresher training was conducted after nine months of implementation, halfway through the intervention. |
| **How** | Home visits using the following techniques knowledge sharing, counselling and problem solving, and opportunities for caregivers to try play and communication activities with their child with feedback and guidance on responsiveness during the interaction. |
| **When and How much** | Every 4-6 weeks with an average duration of 35 minutes. The program was delivered for 18 months. |
| **Tailoring** | The responsive stimulation component was a Tanzanian and Swahili adapted version of the UNICEF and WHO Care for Development package. Stimulation components were tailored by child age and abilities. |
| **How well** | The intervention was delivered with high fidelity (See Table 2 for indicators) |

**Supplemental Table 2.** Mean composite Bayley Scales of Infant and Toddler Development–III scores at 18 months of follow-up by nurse assessor

|  | **Assessor #1**  **(n=195)** | **Assessor #2**  **(n=210)** |
| --- | --- | --- |
| Cognitive composite score | 92.4±11.5 | 97.4±9.0 |
| Language composite score | 89.1±9.2 | 102.66±10.3 |
| Motor composite score | 97.7±10.7 | 104.79±11.0 |

**Supplemental Table 3.** Internal consistency as measured by Cronbach’s alpha for Bayley Scales of Infant and Toddler Development Scores –III raw scores in the full sample and by nurse assessor

|  | **Internal consistency**  **(Cronbach’s alpha)** | | |
| --- | --- | --- | --- |
|  | **Full Sample**  **(n=405)** | **Assessor #1**  **(n=195)** | **Assessor #2**  **(n=210)** |
| Cognitive | 0.92 | 0.91 | 0.93 |
| Language | 0.94 | 0.94 | 0.94 |
| Motor | 0.94 | 0.94 | 0.94 |

**Supplemental Table 4.** Assessment of potential imbalance of baseline characteristics between trial arms

|  | **Community Health Worker Mean ± SD or N (%)** | **Community Health Worker + Conditional Cash Transfer Mean ± SD or N (%)** | **Control Mean ± SD or N (%)** | **p-value** |
| --- | --- | --- | --- | --- |
| *Household characteristics* |  |  |  |  |
| Household size (persons) | 4.0 ± 1.7 | 3.6 ± 2.0 | 3.4 ± 1.7 | 0.24 |
| Improved sanitation | 175 (87.5%) | 103 (51.5%) | 123 (63.7%) | 0.08 |
| Wealth quintile | (15%) |  |  |  |
| Q1 – Poorest | 29 (14.5%) | 50 (25.0%) | 44 (22.8%) | <0.01 |
| Q2 | 40 (20.0%) | 57 (28.5%) | 23 (11.9%) |  |
| Q3 | 42 (21.0%) | 49 (24.5%) | 34 (17.6%) |  |
| Q4 | 40 (20.0%) | 30 (15.0%) | 47 (24.4%) |  |
| Q5 - Richest | 49 (24.5%) | 14 (7.0%) | 45 (23.3%) |  |
| *Maternal characteristics* |  |  |  |  |
| Age, years | 26.9 ± 5.3 | 27.0 ± 6.3 | 26.4 ± 6.1 | 0.24 |
| Married or living with partner | 172 (86.0%) | 167 (83.5%) | 149 (77.2%) | 0.48 |
| Education |  |  |  | 0.09 |
| No formal education | 8 (4.0%) | 36 (18.0%) | 8 (4.2%) |  |
| Primary education | 179 (89.5%) | 139 (69.5%) | 155 (80.3%) |  |
| Secondary or higher education | 13 (6.5%) | 25 (12.5%) | 30 (15.5%) |  |
| Pregnant at time of enrollment | 55 (27.5%) | 75 (37.5%) | 67 (34.7%) | 0.23 |
| Multiparous | 187 (93.5%) | 179 (89.5%) | 151 (78.2%) | 0.01 |
| High social support (>median) | 55 (27.5%) | 153 (76.5%) | 102 (52.9%) | 0.02 |
| High Caregiver Knowledge of Child Development Inventory score (> median) | 132 (66%) | 80 (40%) | 102 (52.9%) | 0.01 |
| *Infant characteristics* |  |  |  |  |
| Infants (0-1 year at enrollment) | 145 | 125 | 125 |  |
| Male | 75 (51.7%) | 70 (56.0%) | 62 (49.6%) | 0.46 |
| Age, months | 5.3±3.6 | 5.0±3.5 | 4.6±3.1 | 0.23 |

|  | **Child had endline anthropometric**  **data**  **(n=547)** | **Child did not have endline anthropometric**  **data**  **(n=46)** | **p-value** |
| --- | --- | --- | --- |
| Household size | 3.7±1.8 | 3.5±1.9 | 0.52 |
| House has dirt floor | 274 (50.4%) | 22 (47.8%) | 0.72 |
| Household has an improved latrine | 369 (67.5%) | 32 (69.6%) | 0.62 |
| Poorest wealth quintile | 111 (20.4%) | 12 (26.1%) | 0.31 |
| At least 1 toy in the home | 36 (10.8%) | 0 (0%) | 0.06 |
| Mother/caregiver age (in years) | 26.8±6.1 | 26.2±2.1 | 0.08 |
| Mother is married or lives with partner | 453 (82.8%) | 35 (76.1%) | 0.35 |
| Maternal education |  |  |  |
| No formal education | 49 (9.0%) | 3 (6.5%) | 0.70 |
| Primary education | 436 (79.7%) | 37 (80.4%) |  |
| Secondary or higher education | 62 (11.3%) | 6 (13.0%) |  |
| Pregnant at baseline | 172 (31.4%) | 25 (54.4%) | 0.06 |
| Multiparous mother | 481 (87.9%) | 36 (78.3%) | 0.05 |
| Depression HSCL-8 >=1.06 | 321 (58.7%) | 25 (54.4%) | 0.53 |
| Social support scale (1-4) | 2.6±0.8 | 2.7±0.8 | 0.88 |
| Number of stimulation activities reported (0-6) | 1.4±1.3 | 0.8±1.2 | 0.03 |
| CKCDI (0-40) | 15.7±5 | 14.5±5.9 | 0.22 |

**Supplemental Table 5.** Baseline characteristics of children who had endline anthropometric data as compared to children that did not have endline data

**Supplemental Table 6.** Baseline characteristics of children who had endline child development data as compared to children that did not have endline data

|  | **Child had endline development**  **data**  **(n=405)** | **Child did not have endline development**  **data**  **(n=187)** | **p-value** |
| --- | --- | --- | --- |
| Household size | 3.7±1.8 | 3.6±1.8 | 0.37 |
| House has dirt floor | 197 (48.88%) | 99 (52.94%) | 0.43 |
| Household has an improved latrine | 274 (67.65%) | 127 (67.55%) | 0.98 |
| Poorest wealth quintile | 85 (21.09%) | 38 (20.32%) | 0.84 |
| At least 1 toy in the home | 26 (10.4%) | 10 (9.8%) | 0.86 |
| Mother/caregiver age (in years) | 27±6.4 | 26.3±4.6 | 0.12 |
| Mother is married or lives with partner | 339 (83.7%) | 149 (79.26%) | 0.27 |
| Maternal; education |  |  |  |
| No formal education | 30 (7.41%) | 22 (11.7%) | 0.84 |
| Primary education | 333 (82.22%) | 140 (74.47%) |  |
| Secondary or higher education | 42 (10.37%) | 26 (13.83%) |  |
| Pregnant woman | 120 (29.63%) | 77 (40.96%) | 0.08 |
| Multiparous mother | 360 (88.89%) | 157 (83.51%) | 0.05 |
| Depression HSCL-8 >=1.06 | 227 (56.05%) | 119 (63.3%) | 0.04 |
| Social support scale (1-4) | 2.7±0.8 | 2.6±0.8 | 0.14 |
| Number of stimulation activities reported (0-6) | 1.5±1.3 | 1.2±1.2 | 0.05 |
| CKCDI (0-40) | 15.8±5 | 15.1±5.2 | 0.35 |

**Supplemental Table 7.** Effect of CHW + CCT intervention on monthly child health and growth monitoring clinic visit attendance

|  | **CHW+CCT (n=188) Mean ± SD** | **CHW (n=185) Mean ± SD** | **Control (n=174) Mean ± SD** | **CHW+CCT vs. Control Mean Difference (95% CI)** | **CHW vs. Control Mean Difference (95% CI)** |
| --- | --- | --- | --- | --- | --- |
| Number of child health visits attended | 16.1 ± 3.2 | 14.6 ± 4.2 | 13.0 ± 4.7 | 3.0 (2.1-4.0) | 1.5 (0.6-2.5) |

**Supplemental Table 8.** Effect of CHW and CHW+CCT arms on Bayley Scales of Infant and Toddler Development Scores –III composite scores at 18 months of follow-up

|  | | | | **Primary minimally adjusted analysis*** | | **Multivariable adjusted analysis**** | |
| --- | --- | --- | --- | --- | --- | --- | --- |
|  | **CHW Mean ± SD**  **(N=136)** | **CHW+CCT Mean ± SD**  **(N=135)** | **Control Mean ± SD**  **(N=134)** | **CHW vs. Control Mean Difference (95% CI)** | **CHW+CCT vs. Control Mean Difference (95% CI)** | **CHW vs. Control Mean Difference (95% CI)** | **CHW+CCT vs. Control Mean Difference (95% CI)** |
| Cognitive | 95.0 ± 10.2 | 94.8 ± 10.9 | 95.2 ± 10.6 | 2.4 (0.4, 4.5) | 3.4 (1.7, 5.1) | 2.3 (0.6, 3.9) | 3.2 (2.0, 4.5) |
| Language | 96.0 ± 11.5 | 93.5 ± 11.1 | 99.0 ± 12.5 | 0.8 (-1.0, 2.5) | 2.0 (0.9, 3.0) | 0.7 (-0.7, 2.2) | 2.1 (0.2, 3.9) |
| Motor | 100.7 ± 12.1 | 101.2 ±10.9 | 102.3 ±11.2 | 0.8 (-2.2, 3.8) | 3.3 (0.3, 6.4) | 0.9 (-2.0, 3.7) | 3.8 (0.4, 7.1) |

*Minimally adjusted model included covariates for child age at assessment, sex, and BSID-III assessor and accounted for clustering

**Multivariate model included covariates for urban/rural residence, baseline household wealth quintile, household having access to an improved latrine, maternal education, parity, social support, CKCDI, child sex, child age at assessment, sex, and BSID-III assessor and accounted for clustering

**Supplemental Table 9.** Effect of CHW and CHW+CCT arms on standardized mean difference in Bayley Scales of Infant Development Scores –III at 18 months of follow-up using stabilized censoring weights to account for dependent censoring (i.e. loss-to-follow-up)

|  | **Primary minimally adjusted analysis with inverse probability weights for censoring*** | | **Multivariable adjusted analysis with inverse probability weights for censoring**** | |
| --- | --- | --- | --- | --- |
|  | **CHW vs. Control Mean Difference (95% CI)** | **CHW+CCT vs. Control Mean Difference (95% CI)** | **CHW vs. Control Mean Difference (95% CI)** | **CHW+CCT vs. Control Mean Difference (95% CI)** |
| Cognitive | 0.20 (0.12, 0.29) | 0.19 (0.08, 0.30) | 0.16 (0.03, 0.28) | 0.15 (0.07, 0.24) |
| Language | 0.06 (-0.04, 0.16) | 0.08 (-0.01, 0.18) | 0.05 (-0.07, 0.17) | 0.10 (-0.02, 0.23) |
| Motor | 0.09 (-0.02, 0.20) | 0.17 (0.02, 0.32) | 0.07 (-0.05, 0.18) | 0.19 (0.00, 0.38) |

*Minimally adjusted model included covariates for, child age at assessment, sex, and BSID-III assessor and accounted for clustering and used stabilized censoring weights to account for dependent censoring

**Multivariate model included covariates for urban/rural residence, baseline household wealth quintile, household having access to an improved latrine, maternal education, parity, social support, CKCDI, child sex, child age at assessment, sex, and BSID-III assessor and accounted for clustering and used stabilized censoring weights to account for dependent censoring

**Supplemental Table 10.** Effect of CHW and CHW + CCT arms collapsed on child development domain z-scores

|  | **CHW and CHW+CCT Combined Mean ± SD**  **(N=271)** | **Control Mean ± SD**  **(N=134)** | **Minimally adjusted***  **Mean Difference (95% CI)** | **Multivariable adjusted****  **Mean Difference (95% CI)** |
| --- | --- | --- | --- | --- |
| Cognitive | 0.05±0.99 | -0.10±1.01 | 0.16 (0.08, 0.24) | 0.14 (0.07, 0.21) |
| Language | -0.03±1.00 | 0.06±1.00 | 0.06 (-0.02, 0.14) | 0.05 (-0.04, 0.13) |
| Motor | 0.03±0.99 | -0.07±1.02 | 0.09 (-0.02, 0.20) | 0.09 (-0.05, 0.23) |

*Minimally adjusted model included covariates for, child age at assessment, sex, and BSID-III assessor and accounted for clustering

**Multivariate model included covariates for urban/rural residence, baseline household wealth quintile, household having access to an improved latrine, maternal education, parity, social support, CKCDI, child sex, child age at assessment, sex, and BSID-III assessor and accounted for clustering

**Supplemental Table 11**. Effect modification of randomized arm on development outcomes by pre-defined factors, multivariable adjusted

|  | **Cognitive domain z-score*** | | **Language domain z-score*** | | **Motor domain z-score*** | |
| --- | --- | --- | --- | --- | --- | --- |
|  | **CHW vs. Control Mean Difference (95% CI)** | **CHW+CCT vs. Control Mean Difference (95% CI)** | **CHW vs. Control Mean Difference (95% CI)** | **CHW+CCT vs. Control Mean Difference (95% CI)** | **CHW vs. Control Mean Difference (95% CI)** | **CHW+CCT vs. Control Mean Difference (95% CI)** |
| Pregnancy status at trial enrollment |  |  |  |  |  |  |
| Not pregnant – Child < 1 year | 0.12 (0.01, 0.22) | 0.08 (-0.01, 0.18) | -0.03 (-0.14, 0.09) | 0.01 (-0.04, 0.06) | 0.08 (-0.06, 0.21) | 0.21 (0.00, 0.42) |
| Pregnant | 0.16 (-0.13, 0.44) | 0.26 (0.03, 0.49) | 0.16 (-0.09, 0.40) | 0.19 (-0.09, 0.47) | -0.07 (-0.30, 0.16) | 0.1 (-0.08, 0.27) |
| p-value for interaction | 0.82 | 0.26 | 0.23 | 0.21 | 0.24 | 0.29 |
| Maternal education |  |  |  |  |  |  |
| Less than secondary school | 0.12 (0.04, 0.20) | 0.12 (0.05, 0.18) | 0.02 (-0.09, 0.12) | 0.06 (-0.03, 0.16) | 0.05 (-0.08, 0.18) | 0.21 (0.05, 0.38) |
| Secondary school or greater | 0.19 (-0.01, 0.4) | 0.35 (0.00, 0.70) | 0.17 (-0.05, 0.39) | 0.15 (-0.11, 0.41) | 0.05 (-0.07, 0.17) | -0.11 (-0.39, 0.16) |
| p-value for interaction | 0.42 | 0.20 | 0.24 | 0.55 | 0.98 | 0.01 |
| Maternal age |  |  |  |  |  |  |
| <25 years | 0.1 (-0.08, 0.27) | 0.14 (0.01, 0.28) | 0.06 (-0.06, 0.18) | 0.04 (-0.08, 0.17) | 0.07 (-0.07, 0.21) | 0.16 (-0.02, 0.34) |
| ≥25 years | 0.21 (0.06, 0.37) | 0.15 (-0.02, 0.33) | 0.00 (-0.13, 0.14) | 0.13 (-0.02, 0.28) | 0.03 (-0.23, 0.29) | 0.24 (-0.04, 0.52) |
| p-value for interaction | 0.44 | 0.93 | 0.55 | 0.40 | 0.77 | 0.55 |
| Household wealth |  |  |  |  |  |  |
| <50^th^ percentile | 0.17 (0.06, 0.29) | 0.22 (0.09, 0.35) | 0.04 (-0.07, 0.14) | 0.00 (-0.15, 0.15) | 0.09 (-0.13, 0.31) | 0.24 (-0.03, 0.52) |
| ≥50^th^ percentile | 0.1 (-0.01, 0.21) | 0.05 (-0.13, 0.24) | 0.03 (-0.11, 0.17) | 0.17 (0.02, 0.32) | -0.01 (-0.1, 0.08) | 0.12 (0.01, 0.22) |
| p-value for interaction | 0.23 | 0.25 | 0.98 | 0.19 | 0.36 | 0.33 |
| Maternal depression |  |  |  |  |  |  |
| No | 0.18 (0.06, 0.30) | 0.14 (-0.03, 0.32) | 0.06 (-0.03, 0.15) | 0.08 (-0.06, 0.21) | 0.13 (-0.02, 0.28) | 0.22 (0.03, 0.42) |
| Yes (HSCL-8 ≥1.06) | 0.08 (-0.10, 0.26) | 0.13 (-0.03, 0.29) | 0.00 (-0.21, 0.21) | 0.06 (-0.09, 0.22) | -0.04 (-0.24, 0.16) | 0.15 (-0.06, 0.35) |
| p-value for interaction | 0.47 | 0.93 | 0.64 | 0.91 | 0.22 | 0.54 |
| Social support |  |  |  |  |  |  |
| <50^th^ percentile | 0.15 (0.06, 0.24) | 0.16 (0.03, 0.28) | -0.04 (-0.18, 0.10) | -0.01 (-0.24, 0.22) | 0.03 (-0.06, 0.13) | 0.2 (-0.11, 0.51) |
| ≥50^th^ percentile | 0.12 (-0.05, 0.29) | 0.13 (0.01, 0.25) | 0.11 (0.00, 0.22) | 0.13 (0.00, 0.26) | 0.06 (-0.08, 0.20) | 0.17 (-0.01, 0.35) |
| p-value for interaction | 0.77 | 0.82 | 0.069 | 0.38 | 0.62 | 0.87 |
| Maternal knowledge of child development |  |  |  |  |  |  |
| Lower CKCDI scores (< 50 percentile) | 0.18 (-0.10, 0.46) | 0.19 (0.06, 0.32) | 0.16 (-0.07, 0.39) | 0.18 (0.06, 0.30) | 0.10 (-0.24, 0.44) | 0.25 (0.02, 0.47) |
| Higher CKCDI scores (≥50^th^ percentile) | 0.10 (-0.04, 0.25) | 0.09 (-0.07, 0.25) | -0.05 (-0.16, 0.05) | -0.04 (-0.16, 0.08) | 0.00 (-0.16, 0.16) | 0.11 (-0.14, 0.36) |
| p-value for interaction | 0.70 | 0.45 | 0.11 | 0.01 | 0.68 | 0.41 |

*Multivariate model included covariates for urban/rural residence, baseline household wealth quintile, household having access to an improved latrine, maternal education, parity, social support, CKCDI, child sex, child age at assessment, sex, and BSID-III assessor and accounted for clustering

**Supplemental Table 12.** Effect of CHW and CHW+CCT arms on child length/height-for-age z-scores (LAZ/HAZ), weight-for-length/height z-scores (WLZ/WHZ) and weight-for-age z-scores using stabilized censoring weights to account for dependent censoring (i.e. loss-to-follow-up)

|  | **Primary minimally adjusted analysis with inverse probability weights for censoring*** | | **Multivariable adjusted analysis with inverse probability weights for censoring**** | |
| --- | --- | --- | --- | --- |
|  | **CHW vs. Control Mean Difference (95% CI)** | **CHW+CCT vs. Control Mean Difference (95% CI)** | **CHW vs. Control Mean Difference (95% CI)** | **CHW+CCT vs. Control Mean Difference (95% CI)** |
| LAZ/HAZ | 0.81 (-0.62, 2.24) | 1.60 (0.14, 3.06) | 0.52 (-0.20, 1.24) | 1.38 (0.55, 2.21) |
| WAZ | 0.13 (-0.17, 0.44) | 0.12 (-0.21, 0.44) | 0.09 (-0.19, 0.37) | 0.04 (-0.31, 0.38) |
| WLZ/WHZ | -0.38 (-1.19, 0.43) | -0.97 (-1.95, 0.00) | -0.24 (-0.63, 0.16) | -0.93 (-1.47, -0.38) |

*Minimally adjusted model included covariates for child age at assessment and sex and accounted for clustering and used stabilized censoring weights to account for dependent censoring

**Multivariate model included covariates for urban/rural residence, baseline household wealth quintile, household having access to an improved latrine, maternal education, parity, social support, CKCDI, child sex, child age at assessment, and sex, and accounted for clustering and used stabilized censoring weights to account for dependent censoring

**Supplemental Table 13.** Effect of CHW and CHW + CCT arms collapsed on child length/height-for-age z-scores (LAZ/HAZ), weight-for-length/height z-scores (WLZ/WHZ) and weight-for-age z-scores

|  | **CHW and CHW+CCT Combined Mean ± SD**  **(N=372)** | **Control Mean ± SD**  **(N=174)** | **Minimally-adjusted* Mean Difference (95% CI)** | **Multivariable* adjusted Mean Difference (95% CI)** |
| --- | --- | --- | --- | --- |
| LAZ/HAZ | -0.56±1.34 | -1.65±1.78 | 1.12 (-0.23, 2.47) | 1.09 (0.56, 1.62) |
| WAZ | -0.03±0.92 | -0.27±0.98 | 0.26 (-0.01, 0.52) | 0.21 (0.00, 0.43) |
| WLZ/WHZ | 0.34±1.28 | 0.78±1.54 | -0.43 (-1.31, 0.45) | -0.46 (-0.87, -0.05) |

*Minimally adjusted model included a covariate for child age at assessment and sex and accounted for clustering

**Multivariate model adjusted included covariates for urban/rural residence (randomization scheme), baseline household wealth quintile, household having access to an improved latrine, maternal education, parity, social support, CKCDI, child sex, and child age at assessment and accounted for clustering

**Supplemental Table 14**. Effect modification of CHW and CHW+CCT interventions on anthropometric outcomes by pre-defined baseline factors, multivariable adjusted

|  | **LAZ/HAZ*** | | **WAZ*** | | **WHZ*** | |
| --- | --- | --- | --- | --- | --- | --- |
|  | **CHW vs. Control Mean Difference (95% CI)** | **CHW+CCT vs. Control Mean Difference (95% CI)** | **CHW vs. Control Mean Difference (95% CI)** | **CHW+CCT vs. Control Mean Difference (95% CI)** | **CHW vs. Control Mean Difference (95% CI)** | **CHW+CCT vs. Control Mean Difference (95% CI)** |
| Pregnancy status at trial enrollment |  |  |  |  |  |  |
| Not pregnant – Child < 1 year | 0.75 (0.01, 1.50) | 1.25 (0.60, 1.90) | 0.21 (-0.03, 0.45) | 0.03 (-0.31, 0.37) | -0.23 (-0.75, 0.29) | -0.85 (-1.37, -0.32) |
| Pregnant | 1.37 (0.57, 2.16) | 1.29 (0.33, 2.25) | 0.37 (-0.11, 0.85) | 0.42 (0.21, 0.63) | -0.37 (-1.25, 0.51) | -0.34 (-1.05, 0.37) |
| p-value for interaction | 0.10 | 0.92 | 0.44 | 0.02 | 0.77 | 0.21 |
| Maternal education |  |  |  |  |  |  |
| Less than secondary school | 1.00 (0.26, 1.74) | 1.33 (0.66, 2.00) | 0.28 (0.01, 0.55) | 0.20 (-0.08, 0.48) | -0.28 (-0.78, 0.22) | -0.67 (-1.19, -0.14) |
| Secondary school or greater | 0.29 (-0.50, 1.07) | 0.80 (0.08, 1.51) | 0.17 (-0.48, 0.81) | -0.10 (-0.73, 0.53) | 0.06 (-1.05, 1.17) | -0.73 (-1.58, 0.12) |
| p-value for interaction | 0.16 | 0.02 | 0.73 | 0.35 | 0.56 | 0.90 |
| Maternal age |  |  |  |  |  |  |
| <25 years | 0.82 (0.10, 1.53) | 1.14 (0.39, 1.90) | 0.07 (-0.22, 0.37) | 0.10 (-0.21, 0.41) | -0.49 (-0.95, -0.02) | -0.69 (-1.2, -0.19) |
| ≥25 years | 1.12 (0.41, 1.82) | 1.50 (0.89, 2.11) | 0.60 (0.25, 0.94) | 0.27 (-0.06, 0.60) | 0.14 (-0.51, 0.79) | -0.67 (-1.23, -0.1) |
| p-value for interaction | 0.11 | 0.12 | <0.01 | 0.35 | 0.01 | 0.91 |
| Household wealth |  |  |  |  |  |  |
| <50^th^ percentile | 1.17 (0.23, 2.12) | 1.59 (0.84, 2.34) | 0.45 (0.12, 0.78) | 0.33 (-0.08, 0.74) | -0.17 (-0.83, 0.48) | -0.69 (-1.33, -0.05) |
| ≥50^th^ percentile | 0.68 (0.2, 1.17) | 0.90 (0.12, 1.69) | 0.09 (-0.14, 0.33) | -0.01 (-0.22, 0.20) | -0.32 (-0.67, 0.03) | -0.64 (-1.03, -0.24) |
| p-value for interaction | 0.17 | 0.11 | 0.01 | 0.04 | 0.45 | 0.83 |
| Maternal depression |  |  |  |  |  |  |
| No | 0.49 (-0.03, 1.00) | 1.14 (0.55, 1.72) | 0.25 (-0.06, 0.57) | 0.13 (-0.22, 0.47) | 0.08 (-0.32, 0.48) | -0.63 (-0.98, -0.29) |
| Yes (HSCL-8 ≥1.06) | 1.32 (0.43, 2.21) | 1.44 (0.65, 2.22) | 0.28 (-0.09, 0.66) | 0.16 (-0.09, 0.41) | -0.55 (-1.11, 0.00) | -0.8 (-1.35, -0.26) |
| p-value for interaction | 0.03 | 0.33 | 0.91 | 0.79 | 0.01 | 0.43 |
| Social support |  |  |  |  |  |  |
| <50^th^ percentile | 1.75 (0.83, 2.67) | 2.32 (1.35, 3.29) | 0.46 (0.06, 0.86) | 0.53 (0.09, 0.96) | -0.54 (-1.33, 0.25) | -0.91 (-1.65, -0.18) |
| ≥50^th^ percentile | 0.11 (-0.14, 0.35) | 0.57 (0.05, 1.09) | 0.11 (-0.09, 0.31) | -0.05 (-0.32, 0.22) | 0.08 (-0.25, 0.41) | -0.49 (-0.88, -0.09) |
| p-value for interaction | <0.01 | <0.01 | 0.13 | 0.02 | 0.13 | 0.25 |
| Maternal knowledge |  |  |  |  |  |  |
| Lower CKCDI scores (< 50 percentile) | 1.15 (0.52, 1.79) | 1.29 (0.57, 2.01) | 0.29 (-0.08, 0.65) | 0.22 (-0.03, 0.48) | -0.37 (-0.85, 0.10) | -0.62 (-1.00, -0.24) |
| Higher CKCDI scores (≥50^th^ percentile) | 0.78 (-0.03, 1.60) | 1.26 (0.52, 2.00) | 0.24 (-0.07, 0.54) | 0.09 (-0.27, 0.45) | -0.20 (-0.80, 0.40) | -0.75 (-1.4, -0.1) |
| p-value for interaction | 0.19 | 0.91 | 0.80 | 0.38 | 0.56 | 0.60 |

*Multivariate model adjusted included covariates for urban/rural residence, baseline household wealth quintile, household having access to an improved latrine, maternal education, parity, social support, CKCDI, child sex, and child age at assessment and accounted for clustering

**Supplemental Table 15.** Effect of the integrated Community Health Worker (CHW) intervention and CHW plus conditional cash transfer (CCT) intervention on anthropometric outcomes at 18 months of follow-up among children who were <1 year at the time of enrollment, including adjustment for baseline values.

|  | | | | **Primary minimally**  **adjusted analysis*** | | **Multivariable adjusted*** | |
| --- | --- | --- | --- | --- | --- | --- | --- |
|  | **CHW Mean ± SD or n (%)**  **N=135** | **CHW+CCT Mean ± SD or n (%)**  **N=122** | **Control Mean ± SD or n (%)**  **N=115** | **CHW vs. Control Mean Difference or Relative Risk (95% CI)** | **CHW+CCT vs. Control Mean Difference or Relative Risk (95% CI)** | **CHW vs. Control Mean Difference or Relative Risk (95% CI)** | **CHW+CCT vs. Control Mean Difference or Relative Risk (95% CI)** |
| Length/Height-for-age z-score (HAZ) | -0.86 ± 1.31 | -0.26 ± 1.31 | -1.65 ± 1.78 | 0.91 (-0.35, 2.18) | 1.58 (0.38, 2.27)^c^ | 0.74 (0.05, 1.42) ^c^ | 1.35 (0.67, 2.03) ^c^ |
| Stunting (HAZ < -2) | 37 (20.2%) | 19 (10.1%) | 66 (38.2%) | 0.54 (0.17, 1.64) | 0.32 (0.11, 0.95) ^c^ | 0.73 (0.38, 1.40) | 0.75 (0.41, 1.38) |
| Weight-for-age z-score (WAZ) | 0.01 ± 0.91 | -0.07 ± 0.93 | -0.27 ± 0.98 | 0.15 (-0.23, 0.54) | 0.10 (-0.40, 0.61) | 0.18 (-0.11, 0.48) | -0.01 (-0.38, 0.36) |
| Underweight (WAZ < -2) | 3 (1.6%) | 5 (2.7%) | 9 (5.2%) | 0.42 (0.09, 1.94) | 0.36 (0.08, 1.61) | 0.14 (0.03, 0.72) ^c^ | 0.00 (0.00, 0.03) ^c^ |
| Weight-for-height z-score (WHZ) | 0.62 ± 1.28 | 0.07 ± 1.23 | 0.78 ± 1.54 | -0.32 (-1.09, 0.46) | -0.92 (-1.83, -0.00)^c^ | -0.20 (-0.82, 0.42) | -0.91 (-1.55, -0.28) ^c^ |
| Wasting (WHZ < -2) | 4 (2.2%) | 10 (5.3%) | 6 (3.5%) | 0.27 (0.03, 2.59) | 0.92 (0.29, 2.88) | 0.07 (0.02, 0.33) ^c^ | 0.57 (0.17, 1.97) |
| Overweight (WHZ > 2) | 28 (15.2%) | 10 (5.3%) | 35 (20.1%) | 0.72 (0.27, 1.88) | 0.32 (0.07, 1.50) | 0.72 (0.37, 1.39) | 0.26 (0.08, 0.85) ^c^ |

^a^Multivariate model included covariates for baseline value of the outcome, urban/rural residence (randomization scheme), baseline household wealth quintile, household having access to an improved latrine, maternal education, parity, social support, CKCDI, child sex, child age, and, and accounted for clustering

^b^ p-value <0.05
